# Proton magnetic resonance spectroscopy in frontotemporal lobar degeneration-related syndromes

**DOI:** 10.1101/2021.01.11.21249589

**Authors:** Alexander G Murley, Kamen A Tsvetanov, Matthew A Rouse, P Simon Jones, Katrine Sværke, Win Li, T Adrian Carpenter, James B Rowe

**Affiliations:** Department of Clinical Neurosciences, University of Cambridge, UK; Cambridge University Hospitals NHS Foundation Trust, UK; MRC Cognition and Brain Sciences Unit, University of Cambridge, UK

**Keywords:** Frontotemporal dementia, corticobasal degeneration, progressive supranuclear palsy, MRS, behavior

## Abstract

**Objective:** To measure cortical metabolite deficits *in vivo* in syndromes associated with frontotemporal lobar degeneration, in relation to cognitive and behavioral change.

**Methods:** Sixty patients were recruited with a clinical syndrome associated with frontotemporal lobar degeneration (behavioral variant frontotemporal dementia n=11, progressive supranuclear palsy n=26, corticobasal syndrome n=11, primary progressive aphasias n=12), and 38 age- and sex-matched healthy controls. We measured nine metabolites in the right inferior frontal gyrus, superior temporal gyrus and right primary visual cortex using 3T semi-laser magnetic resonance spectroscopy. Metabolite concentrations were corrected for age, sex, and partial volume. We related corrected metabolite concentrations to cognitive and behavioral measures using canonical correlation analysis.

**Results:** Metabolite concentrations varied significantly by brain region and diagnosis (region x metabolite x diagnosis interaction F_(64)_=1.73, p<0.001, corrected for age, sex, and atrophy within the voxel). N-acetyl aspartate and glutamate concentrations were reduced in the right prefrontal cortex in behavioral variant frontotemporal dementia and progressive supranuclear palsy, even after partial volume correction. The reduction of these metabolites was associated with executive dysfunction and behavioral impairment (canonical correlation analysis R=0.95, p<0.001).

**Conclusion:** Magnetic resonance spectroscopy confirms behaviourally relevant metabolite deficits including glutamate, in syndromes associated with frontotemporal lobar degeneration. Magnetic resonance spectroscopy may be a useful index of neurodegeneration, and highlight candidates for pharmacological treatment.

## Introduction

Frontotemporal lobar degeneration (FTLD) is a heterogeneous group of diseases characterised by focal neurodegeneration of frontal, temporal and subcortical regions including the basal ganglia and midbrain.^1^ These progressive, incurable diseases cause a spectrum of clinical syndromes; behavioral variant frontotemporal dementia (bvFTD), primary progressive aphasias (PPA), progressive supranuclear palsy (PSP) and corticobasal syndrome (CBS).^2–5^ However the clinical phenotypes associated with FTLD are heterogeneous and overlapping in their features^6^, and include higher cognitive changes which cause a high burden on the patient and their families and carers.^7,8^. There is a pressing need for a better mechanistic understanding of their pathophysiology, in support of new treatment strategies.

Here, using magnetic resonance spectroscopy (MRS) we quantify *in vivo* the biochemical consequences of FTLD. Our aim was to compare metabolite profiles in the frontal and temporal lobes, the cortical regions most typically affected in FTLD, with the occipital lobe that is relatively spared by the diseases. We predefined the right inferior frontal and superior temporal gyri as regions of interest. These regions are affected by multiple FTLD syndromes^1,9^ and form part of a network regulating behavior.^10,11^ We tested the hypothesis that metabolites associated with neuronal structure and function would be reduced in the frontal and temporal lobes in disorders associated with FTLD, and that this metabolite deficit would correlate with the severity of cognitive and behavioral impairment.

## Methods

### Participant recruitment and testing

Participants were recruited as part of the PIPPIN (“Pick’s disease and Progressive supranuclear palsy prevalence and incidence”) study, an epidemiological cohort study of FTLD-related syndromes in the East of England. Details of the study have been reported elsewhere ^6,12,13^. In brief, PIPPIN aimed to recruit all patients living with a FTLD syndrome in the UK counties of Cambridgeshire and Norfolk. All patients met the clinical diagnostic criteria for a principal FTLD syndrome ^2–5^. We grouped all PSP phenotypes into the PSP group, and all progressive aphasia subtypes in to the PPA group, in view of the group size. Control participants with no neurological or psychiatric disease were recruited from the NIHR Join Dementia Research register. Participants were invited for clinical examination, cognitive testing, and MRI. Here, we report the subset of patients (n=60) and age and sex matched healthy controls (n=38) who completed magnetic resonance spectroscopy. All participants provided written informed consent or, if they lacked capacity to consent, their next of kin were consulted using the ‘personal consultee’ process established in UK law. The study had ethical approval from the Cambridge Central Research Ethics Committee (REC 12/EE/0475).

Each participant had a structured clinical examination which recorded the presence or absence of clinical features in the current consensus diagnostic criteria ^2–5^. We grouped these features into behavior, language and sensorimotor “scores” based on the sum of features in each group. The behavior score comprised impulsivity, apathy, loss of empathy, stereotyped and/or compulsive behaviors, hyperorality and dietary change. The language score included agrammatic, apraxic and logopenic speech and impaired semantic memory. The sensorimotor score comprised cortical sensory loss, apraxia and alien limb syndrome, and sensorimotor deficits. Participants completed formal cognitive testing including the Addenbrookes Cognitive Examination – Revised (ACE-R) and the Frontal Assessment Battery (FAB), which is sensitive to executive dysfunction in FTLD syndromes.^14^ Participants’ nearest relative or carer completed the revised Cambridge Behavioural Inventory (CBI-R).

### Magnetic resonance spectroscopy

Participants were scanned at the Wolfson Brain Imaging Centre, University of Cambridge on a Siemens 3T PRISMA system. A T1-weighted structural sequence (MPRAGE TR=2000ms,TE=2.93ms, TI=850ms, FA=8°, 208 slices, 1.1mm isotropic voxels) was acquired for localisation of the spectroscopy regions of interest (ROI) and partial volume correction. Single-voxel magnetic resonance spectra were acquired serially from 2×2×2cm voxels placed manually by the same operator (AGM) over the right inferior frontal gyrus, right superior temporal gyrus and right primary visual cortex using anatomical landmarks. Spectra were acquired using a short-echo semi-LASER sequence (64 repetitions, TR/TE = 5000/28 ms)^15,16^ with FASTESTMAP shimming^17^ and water-peak flip angle and VAPOR water suppression.^18^ The 64 individual repetitions were saved separately then corrected for eddy current effect, frequency and phase shifts using MRspa (Dinesh Deelchand, University of Minnesota, www.cmrr.umn.edu/downloads/mrspa). All spectra were visually inspected for quality control. Spectra from 18 voxels (FTLD n=9, Control n=9) were excluded due to movement artefact, inadequate water suppression and/or lipid contamination.

Neurochemicals between 0.5 and 4.2ppm were quantified using LCModel (Version 6.2-3)^19^ with water scaling and a simulated basis set that included an experimentally-acquired macromolecule spectra. The fractions of grey matter, white matter and CSF were obtained from segmentation of the MP2RAGE imaging using the standard voxel-based morphometry pre-processing pipeline in SPM12. Nine metabolites had a mean Cramer Rao Lower Bound lower than 20 and were saved for further analysis. Metabolites with correlation ≤-0.3 were reported together: (i) NAA and NAAG, (ii) choline and glycerophosphocholine, (iii) creatine and phosphocreatine, (iv) glucose and taurine and (v) ascorbate and glutathione. Measures of scan quality, including water linewidth and signal to noise, are reported in table 1.

**Table 1:** Magnetic resonance spectroscopy quality metrics. Values for control and FTLD-associated syndromes are the group mean (standard deviation in parentheses).CRLB=Cramer Rao Lower Bound. IFG=inferior frontal gyrus, STG=superior temporal gyrus, V1=right primary visual cortex. t=Welch t-test comparing all FTLD syndromes with control, df=degrees of freedom.

| Region | Quality measure | Control | FTLD | t | df | p |
| --- | --- | --- | --- | --- | --- | --- |
| Right IFG | Glutamate CRLB | 3.9 (0.4) | 4.8 (1.6) | -3.7 | 71 | <0.001 |
|  | NAA+NAAG CRLB | 2.0 (0.2) | 2.1 (0.5) | -1.8 | 92.4 | 0.08 |
|  | Water linewidth | 7.3 (0.8) | 6.3 (1.2) | 5.13 | 95.5 | <0.001 |
|  | Signal to noise ratio | 56.4 (5.2) | 47.3 (8.6) | 6.53 | 95.8 | <0.001 |
| Right STG | Glutamate CRLB | 4.3 (1.0) | 4.9 (1.4) | -2.5 | 82 | 0.02 |
|  | NAA+NAAG CRLB | 2.0 (0.3) | 2.1 (0.6) | -1.6 | 76.7 | 0.12 |
|  | Water linewidth | 9.00 (0.9) | 8.4 (1.6) | 1.99 | 77.4 | 0.05 |
|  | Signal to noise ratio | 49.7 (9.3) | 45.3 (11.3) | 1.95 | 81.2 | 0.05 |
| Right V1 | Glutamate CRLB | 5.1 (2.9) | 4.7 (2.8) | 0.64 | 59.5 | 0.53 |
|  | NAA+NAAG CRLB | 1.9 (0.8) | 1.8 (0.7) | 0.62 | 56.6 | 0.54 |
|  | Water linewidth | 7.5 (0.8) | 7.2 (0.8) | 1.38 | 63.2 | 0.17 |
|  | Signal to noise ratio | 58.1 (16.0) | 61.6 (13.8) | -1 | 55.1 | 0.32 |

### Statistical analysis

A generalised linear model was used to remove the effect of age, sex and partial volume (of grey and white matter) from the metabolite concentrations. This model was weighted with the metabolite Cramer Rao Lower Bound and the residuals used for further analysis. The tissue correction method was the same as Murley at et (2020).^20^ Factorial analysis of variance (ANOVA) was used to compare nineteen metabolite concentrations across three brain regions and between five groups. Dunnett’s post hoc test was used to explore differences between control participants and the FTLD syndrome subtypes. Welch’s t-test was used to test for differences in cognitive tests between healthy participants and those with a FTLD syndrome.

Since both cognition and metabolites are multivariate, we used canonical correlation analysis (CCA) to compare the frontal and temporal lobe metabolite profiles (as measured by MRS) and cognitive and behavioral impairment (as measured by the ACE-R, FAB, CBI and clinician rating of behavioral, language and sensorimotor impairment). CCA is a multivariate technique that measures the association between two sets of variables.^21^ It uses a data-driven approach to reveal latent, common factors, or canonical covariates, underlying these associations.^6,22^ All variables were standardised into z-scores before CCA. The CCA was permuted 5000 times to determine significance and ensure stability of the final components.

### Data availability

Anonymised data are available on reasonable request for academic use, subject to restrictions required to protect participant confidentiality.

## Results

Participant characteristics are summarised in Table 2. There was no significant difference in the age and sex distributions of the patient and control groups. Patients had marked global cognitive and behavioral impairment, as rated by direct patient measures (ACE-R, FAB), structured carer interview (CBI-R) and clinician rating based on history and examination (Table 2).

**Table 2:**
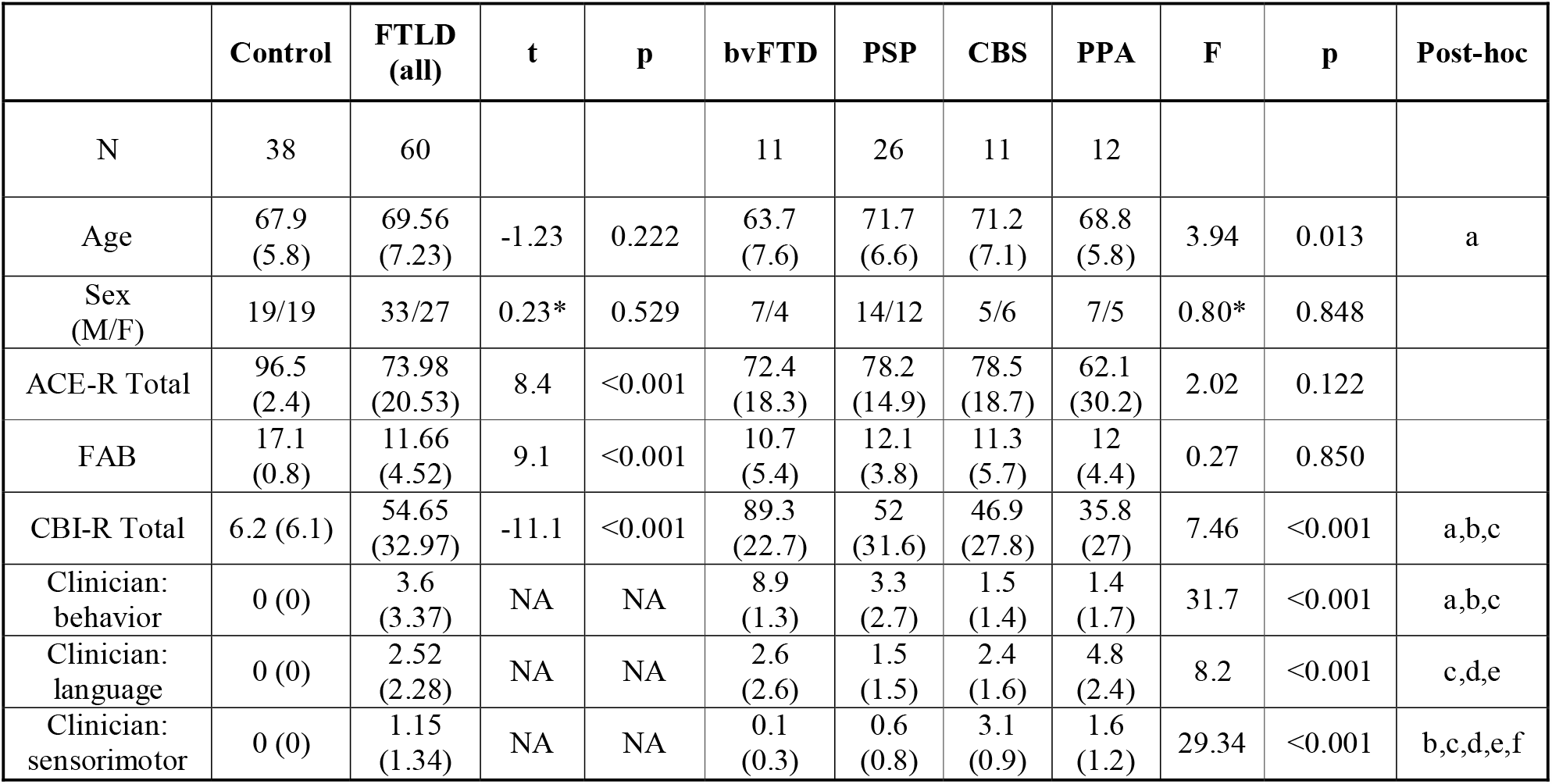
Participant demographics and cognitive test results. FTLD=all FTLD-related syndromes, bvFTD=behavioral variant frontotemporal dementia, PSP=progressive supranuclear palsy, CBS=corticobasal syndrome, PPA=primary progressive aphasia (all subtypes), ACE-R: Addenbrooke’s Cognitive Examination – Revised, FAB: Frontal Assessment Battery, CBI-R: Cambridge Behavioral Inventory – Revised. Clinician behavior, sensorimotor and language are the sum of the presence of features in each domain, t= Welch t-test comparing all FTLD syndromes with control, ^*\**^*=* ^*2*^ test, F=ANOVA of FTLD syndrome subtypes, Post-hoc pairwise Tukey’s tests (p<0.05): a=bvFTD vs PSP, b=bvFTD vs CBS, c=bvFTD vs PPA,d=PPA vs PSP, e=PPA vs CBS, f=CBS vs PSP, NA = no variance in control group.

Our analyses proceeded in two stages. First, we used single-voxel magnetic resonance spectroscopy (MRS) to measure nine metabolites in the frontal, temporal and occipital lobes. Metabolite concentrations, after correction for age, sex, and atrophy within the voxel, varied significantly by region and diagnosis (region x metabolite x diagnosis interaction F_(64)_=1.73, p<0.001).

In other words, there were regionally specific effects of disease, for some but not all metabolites. These can be understood in the context of the first order interactions and main effects (Table 3) as follows. Metabolite concentrations varied by region but there was no region by diagnosis interaction (F_(8)_=1.23 p=0.29), validating the effective partial volume correction of the metabolite concentrations. Main effect analysis showed that the region by metabolite by diagnosis interaction was due to differences in neurotransmitter glutamate and neuronal marker N-acetyl-aspartate and N-acetyl-asparate-glutamate (NAA+NAAG) concentration. *Post hoc* analyses revealed lower concentrations of the neurotransmitter glutamate and neuronal marker N-acetyl aspartate in bvFTD and PSP compared to controls (Figure 1). A glutamate deficit was also found in PSP in the right superior temporal gyrus (Figure 1). N-acetyl aspartate concentrations were low in the right visual cortex in PSP. Despite these significant group differences, there was wide variation in metabolite concentrations in all groups (Figure 1).

**Table 3.** Full factorial analysis of variance showing first order results and simple main effect of diagnosis. Regions: IFG= right inferior frontal gyrus, STG=right superior temporal gyrus, V1=right primary visual cortex. Diagnoses: bvFTD, PSP, CBS, PPA, Control. Metabolites: Aspartate, Glutamine, Glutamate, myo-inositol, N-acetyl aspartate (including N-acetyl-aspartate-glutamate), choline=choline and glycerophosphocholine, creatine= creatine and phosphocreatine

| ANOVA (between and within subject effects) |  |  |  |  |  |
| --- | --- | --- | --- | --- | --- |
| Cases |  | Sum of Squares | df | F | p |
| Region |  | 4.05 | 2 | 4.52 | 0.013 |
| Region x Diagnosis |  | 4.40 | 8 | 1.23 | 0.288 |
| Metabolite |  | 10280.33 | 8 | 2841.54 | <0.001 |
| Metabolite x Diagnosis |  | 31.33 | 32 | 2.17 | <0.001 |
| Region x Metabolite |  | 149.26 | 16 | 55.58 | <0.001 |
| Region x Metabolite x Diagnosis |  | 18.54 | 64 | 1.73 | <0.001 |
| Diagnosis |  | 5.29 | 4 | 0.91 | 0.464 |
| Simple Main Effects - Diagnosis |  |  |  |  |  |
| Level of ROI | Level of Metabolite | Sum of Squares | df | F | p |
| Frontal<br>(Right IFG) | Aspartate | 1.19 | 4 | 1.87 | 0.125 |
|  | Glutamine | 0.80 | 4 | 1.05 | 0.391 |
|  | **Glutamate** | **7.67** | **4** | **5.73** | **<0.001** |
|  | Myoinositol | 2.94 | 4 | 1.41 | 0.240 |
|  | Ascorbate + glutathione | 0.43 | 4 | 1.41 | 0.239 |
|  | Glucose + taurine | 3.87 | 4 | 2.34 | 0.064 |
|  | Choline | 0.10 | 4 | 0.61 | 0.654 |
|  | **NAA + NAAG** | **8.83** | **4** | **4.16** | **0.004** |
|  | Creatine | 0.41 | 4 | 0.63 | 0.646 |
| Temporal<br>(Right STG) | Aspartate | 0.58 | 4 | 0.14 | 0.965 |
|  | Glutamine | 1.19 | 4 | 1.02 | 0.404 |
|  | **Glutamate** | **7.26** | **4** | **2.71** | **0.037** |
|  | Myoinositol | 1.17 | 4 | 0.85 | 0.499 |
|  | Ascorbate + glutathione | 0.40 | 4 | 0.45 | 0.776 |
|  | Glucose + taurine | 1.72 | 4 | 0.98 | 0.426 |
|  | Choline | 0.24 | 4 | 1.52 | 0.206 |
|  | **NAA + NAAG** | **9.84** | **4** | **4.67** | **0.002** |
|  | Creatine | 1.98 | 4 | 2.07 | 0.095 |
| Occipital<br>(Right V1) | Aspartate | 0.94 | 4 | 0.50 | 0.736 |
|  | Glutamine | 0.56 | 4 | 1.26 | 0.295 |
|  | Glutamate | 1.54 | 4 | 1.33 | 0.267 |
|  | Myoinositol | 0.74 | 4 | 0.36 | 0.838 |
|  | Ascorbate + glutathione | 0.18 | 4 | 0.67 | 0.614 |
|  | Glucose + taurine | 1.48 | 4 | 0.64 | 0.634 |
|  | Choline | 0.02 | 4 | 0.33 | 0.860 |
|  | **NAA + NAAG** | **3.15** | **4** | **3.36** | **0.014** |
|  | Creatine | 0.34 | 4 | 0.52 | 0.719 |

**Figure 1:**
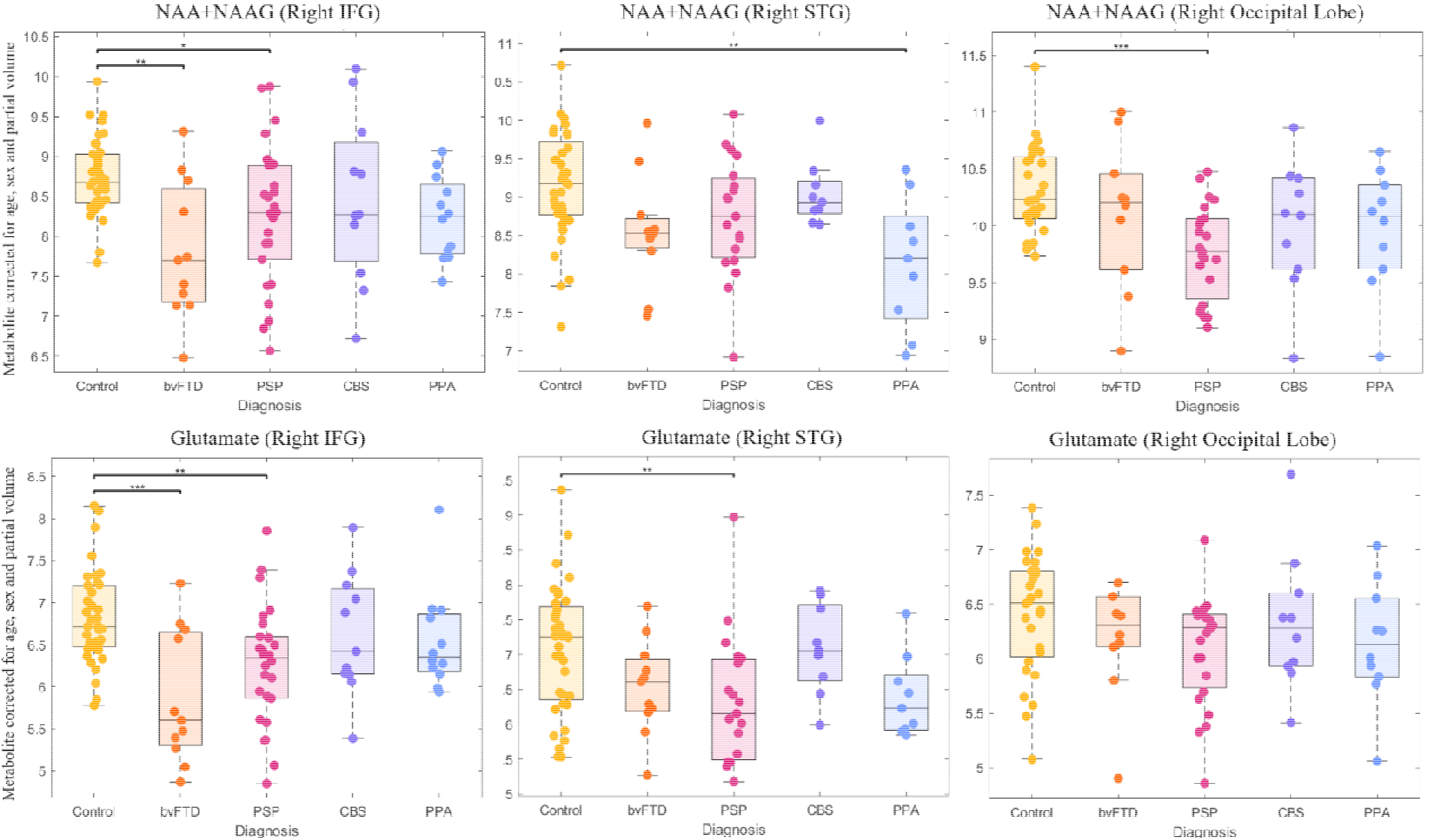
Boxplots of MRS metabolites. Boxplots of metabolites that are significantly different between FTLD-related syndromes and control participants after post-hoc testing from ANOVA of all metabolites, regions and diagnoses. Metabolite values are corrected for age, sex and partial volume. ^*^p<0.05,^**^p<0.01,^***^p<0.001

Correcting for the brain volume differences between groups, had a significant effect on the variation in metabolite concentrations (Factorial ANOVA including corrected and uncorrected data: region x metabolite x uncorrected/corrected x diagnosis F_(64)_=7.94 p<0.001).

Second, we used canonical correlation analysis to test the association between (i) frontal and temporal lobe metabolites and (ii) behavioral and cognitive impairments. This revealed one significant component (R=0.85, p<0.001). This component indicated an association between low glutamate and N-acetyl aspartate in the right inferior frontal gyrus (Figure 2A) and the combination of executive dysfunction, low verbal fluency, low frontal assessment battery scores, and high clinician and carer rating of behavioral impairment (Figure 2D). All FTLD-associated syndromes had strong positive correlations in this component (Figure 2B), and this association was stronger in patients than controls (Figure 2B). There was second component with a trend (p=0.037, not surviving correction for multiple comparisons) with a negative loading from right superior temporal gyrus glutamate, and correlation with carer ratings of impaired everyday skills and abnormal behavior).

**Figure 2:**
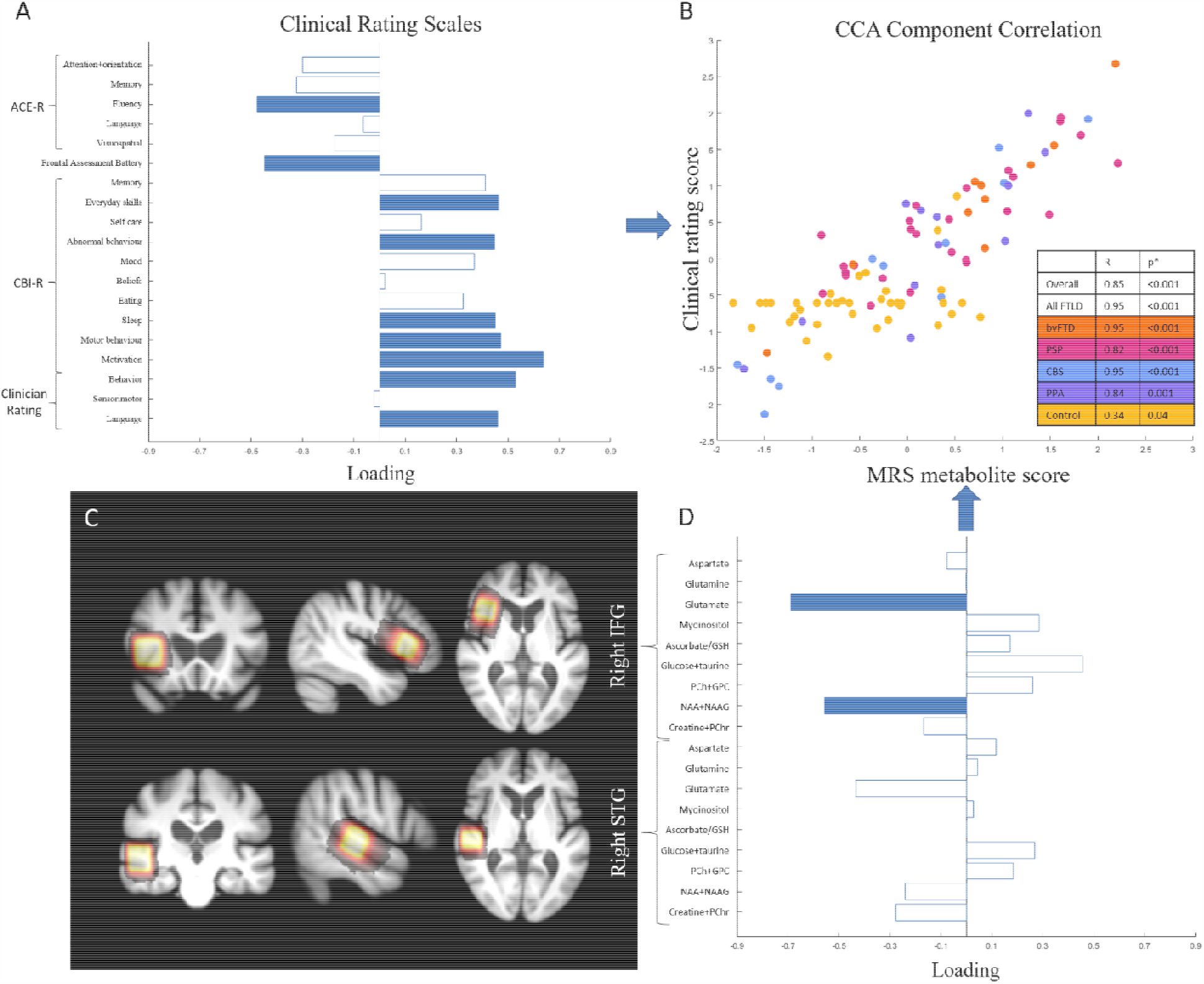
Canonical correlation analysis of MRS and cognitive and behavioral measures. First component from canonical correlation analysis of MRS metabolites and cognitive and behavioral measures. 2A: Loadings from cognitive measures on the first CCA component. Bars colored blue have statistically significant loadings (FWE p<0.05) after permutation testing. Worse performance indicated by Negative ACE-R, positive CBI-R and clinician rating indicate worse cognition/behavior. 2B: First CCA component correlation, color-coded by group. 2C: Map of all participants’ MRS ROIs superimposed on a mean structural image. 2D: MRS metabolite loadings onto the first CCA component. GSH:glutathione, PCh: phosphocholine, GPC: glycerophosphocholine, NAA: N-acetyl-aspartate, NAAG: N-acetyl-aspartate-glutamate, PChr: phosphocreatine Bars colored blue have statistically significant loadings (FWE p<0.05) after permutation testing.

## Discussion

There are two principal results of this study: (i) that N-acetyl aspartate and glutamate are reduced in the prefrontal cortex of people with diverse syndromes associated with frontotemporal lobar degeneration; and (ii) these metabolite differences, as measured by *in vivo* magnetic resonance spectroscopy, correlate with the severity of cognitive and behavioral impairments. A group-wise deficit in these metabolites was seen in bvFTD and PSP, but the association with cognitive and behavioral impairment was in found every patient group. These findings have two implications. First, they strengthen the evidence that metabolite and neurotransmitter deficits are a promising treatment target for pharmacological amelioration of clinical features. Second, because the metabolite deficits in brain tissue are identified after stringent atrophy correction, and correlate with cognition even in syndromes like PSP in which lateral prefrontal cortical atrophy is not marked, the spectroscopy might detect early, even pre-symptomatic, disease before patients develop brain atrophy.

The hypotheses of this study were based on *post mortem* neurochemistry and pre-clinical evidence^23,24^, supported by later ultra-high field MRS.^20,25^ We predicted an association in all FTLD-associated syndromes between glutamate concentration and clinically relevant carer- and clinician-measures of behavioral impairment. Pharmacological correction of such neurochemical deficits might be a tractable target for symptom treatment, especially where these have neurotransmitter functions in addition to metabolic roles. Such symptomatic treatment is a priority given the severe sequelae of cognitive impairment in FTLD.^7,23,26^ However, it remains unclear to what extent our findings reflect a potentially reversible deficit of synaptic glutamate. MRS measures the total pool of unbound glutamate, involved in neuron and glia metabolism, protein synthesis and neurotransmission.^27^ The NMDA antagonist memantine has been used commonly off-licence in frontotemporal dementia treatments, but this is ineffective and anecdotally can worsen cognition in some patients.^28^ Further work is therefore required to understand the relationship between synaptic loss in syndromes associated with FTLD and glutamate deficits.^29^ Healthy age-matched participants also had a weak but statistically significant correlation on this component, corroborating with previous research on glutamate deficits in healthy ageing.^30^

N-acetyl aspartate (NAA) is an abundant amino acid in the central nervous system and comprises the largest peak in the proton spectroscopy spectrum.^31^ NAA is concentrated in neurons and is proposed as a marker of neuronal density, health and function.^31^ Deficits are found in a wide range of neurological diseases associated with neuronal loss including Alzheimer’s disease^32^, stroke^33^ and traumatic brain injury.^34^ NAA is therefore not specific to the proteinopathies associated with FTLD^25^. However, our findings replicate previous studies in FTLD-related syndromes^35–40^ and suggest that NAA levels are a sensitive measure of neuronal loss, over and above structural MRI estimates of brain atrophy. As part of a multi-model MRI battery, NAA spectroscopy may be a useful endpoint in experimental medicine studies, and to understand phenotypic heterogeneity.^41^

We did not replicate the previous studies that reported reduced choline^45^ and elevated *myo*-inositol^45,46^, despite a relatively large sample size, high field strength and use of a consensus sequence and analysis pipelines.^47^ We did find that different partial volume correction methods change the conclusions regarding metabolite concentrations. It is therefore possible that our results are overly stringent in partial volume correction. This has important implications for the clinical use of MRS as some correction methods, e.g. creatine ratios, may be less accurate in measuring metabolite levels in residual brain tissue.

Canonical correlation analysis indicated that glutamate and NAA concentrations in the right inferior frontal gyrus were associated with executive dysfunction and behavioral impairment. The first canonical correlate represented concordant neuropsychological, carer and clinician ratings. This emphasises that frontal NAA and glutamate deficits are associated with clinically relevant cognitive impairment, building on earlier correlations with specific neuropsychological tasks.^20^

Metabolites with lower loadings on the first canonical correlate were not statistically significant, as estimated by permutation testing, but may still be of interest. For example, *myo*-inositol, which is concentrated in glia and elevated with neuroinflammation, had a positive loading.

Neuroinflammation has been identified by TSPO-ligand position emission tomography, in bvFTD, PPA and PSP^42,43^, where it is not only elevated but also prognostic of a more rapid decline.^44^

Our results are relevant to the nosology of syndromes associated with FTLD. The diseases of bvFTD, PSP, CBS, PPA are clinically and pathologically distinct in their classical phenotypes, and there are critical differences in underlying neuropathology even where there is tauopathy. We therefore used the current consensus diagnostic criteria for each clinical disorder. However, the clinical phenotypes associated with FTLD do not respect the diagnostic boundaries and many patients develop clinical features that would meet criteria for more than one disorder.^6^ We have proposed an alternative, *transdiagnostic*, approach to encompass clinical heterogeneity and phenotypic overlap.^6^ This approach, emphasising commonalities in clinic-pathological correlations across the spectrum of FTLD, is supported by our data. The association between prefrontal glutamate, NAA and cognition was observed in all FTLD syndromes jointly, and individually. This is consistent with the metabolite deficits being down-stream of the disease-specific causes of brain injury, and more proximate to the clinical phenotype.

Our study has several limitations. First, this study omits to report GABA concentrations, despite preclinical and ultra-high field MRS evidence of GABA deficits in bvFTD and PSP ^20,23–25^. This omission is because of the low sensitivity and inadequate spectral resolution for GABA using semi-LASER sequence at 3T. Second, the deficits identified by magnetic resonance spectroscopy are unlikely to be specific to one proteinopathy. Participants were diagnosed according to the clinical diagnostic criteria^2–5^ which have variable clinicopathological correlation.^48,49^ In particular, bvFTD is associated with either 3R or 4R tau or TDP-43 pathology, whereas PSP has a high clinicopathological correlation with 4R tau. Our cohort has limited pathological confirmation of the diagnosis but clinicopathological correlations in the PIPPIN study as whole match those found elsewhere.^6,7^ Therefore, while spectroscopy may be a valuable measure of early disease and/or disease progression it is unlikely to differentiate FTLD syndromes according to their underlying proteinopathies. Third, due to small numbers we grouped non-fluent, semantic and logopenic variants of PPA together, although these sub-groups have different clinical and neuropathological features. Fourth, MRS accuracy can be affected by participant movement and other factors which may be greater in the FTLD cohort. To mitigate this we used a consensus guideline-recommended sequence and analysis pipeline^47^ and a within-participant control region in the occipital lobe. Finally, our findings are limited to the single brain regions we imaged. Advances in whole brain MRSI sequences may allow simultaneous measurement of metabolites in multiple regions, better accounting for the clinical and neuropathological heterogeneity in FTLD.

In conclusion, N-acetyl aspartate and glutamate deficits in the prefrontal cortex are associated with loss of executive function and behavioral impairment in each of the major syndromes associated with frontotemporal lobar degeneration (bvFTD, PSP, CBS, PPA), even after correction for atrophy. Magnetic resonance spectroscopy can detect clinically relevant differences in metabolites *in vivo* and may be a valuable adjunct to multi-modal imaging for detecting disease in early stage disease, monitoring progression and response to disease-modifying treatment, and stratification for experimental studies of restorative pharmacology.

## Acknowledgements

We thank the study participants and their families and carers, the radiographers at the Wolfson Brain Imaging Centre, University of Cambridge and Dr Dinesh Deelchand, University of Minnesota, for his MRSpa software (https://www.cmrr.umn.edu/downloads/mrspa/) and MRS basis set.

